# Prevalence of COVID-19 in Adolescents and Youth Compared with Older Adults in States Experiencing Surges

**DOI:** 10.1101/2020.10.20.20215541

**Authors:** Barbara T. Rumain, Moshe Schneiderman, Allan Geliebter

**Affiliations:** Department of Pediatrics, New York Medical College, Valhalla, New York, USA; Department of Psychology, Touro College & University System, New York, N.Y., USA; SUNY Downstate College of Medicine, Brooklyn, New York, USA; Department of Psychiatry, Icahn School of Medicine at Mount Sinai, New York, N.Y., USA

**Author notes:** Address correspondence to: Barbara T. Rumain, Ph.D., Department of Pediatrics, New York Medical College, 40 Sunshine Cottage Road, Valhalla, New York 10595.

## Abstract

**PURPOSE:** There has been considerable controversy regarding susceptibility of adolescents (10-19 years) and youth (15-24 years) to COVID-19. However, a number of studies have reported that adolescents are significantly less susceptible than older adults. Summer 2020 provided an opportunity to examine data on prevalence since after months of lockdowns, with the easing of restrictions, people were mingling, leading to surges in cases.

**METHODS:** We examined data from six U.S. states experiencing surges in the number of cases to determine prevalence of COVID-19, and two other measures, related to prevalence in adolescents and youth as compared to older adults. The two other measures were: (Percentage of cases observed in a given age group) ÷ (percentage of cases expected based on population demographics); and percentage deviation, or [(% observed - % expected)/ % expected] x 100.

**RESULTS:** Prevalence of COVID-19 for adolescents and for youth was significantly greater than for older adults (p<.00001), as was percentage observed ÷ percentage expected (p<.005). The percentage deviation was significantly greater in adolescents/youth than in older adults (p < 0.00001) when there was an excess of observed cases over what was expected, and significantly less when observed cases were fewer than expected (p< 0.00001).

**CONCLUSIONS:** Our results are contrary to previous findings that adolescents are less susceptible than older adults. The findings have implications for school re-openings. The age groups 10-19 and 15-24 are students in middle school, high school, college, and the first two years of professional/graduate school. The high prevalence in these age groups would argue against school re-openings in the near future.

## INTRODUCTION

There has been considerable controversy regarding susceptibility of adolescents and youth to COVID-19. As per the WHO, adolescents are defined to be ages 10-19, and youth are 15-24 years of age. In the very early studies in China, Dong et al.^1^ and Lu et al.^2^ reported that adolescents were susceptible. Bi *et al*., studied individuals from birth to 70+ in China and found the rate of infection in all age groups was similar.^3^ In contrast, Zhang et al., studying individuals in Hunan province, China, concluded that the infection rate in 0-14-year-olds, as reported by the Hunan CDC, was 6.2% as compared to 8.6% among those 15-64 years and 16.3% among those 65 years and above, and that these differences were significant.^4^ Hence, older adults were deemed to be the most susceptible, and those in the first half of adolescence least susceptible, and youth (15-24) were intermediate in susceptibility. Geliebter, Rumain & Schneiderman attempted to replicate Zhang *et al*.’s statistical analyses and obtained results in line with those of Bi *et al*., indicating a similar infection rate for all the age groups.^5^ In Europe, Kuchar et al. found that adolescents were less prone to SARS-CoV-2 infection than adults during the COVID-19 pandemic in Warsaw.^6^ Similarly, in a large cross-sectional analysis using data from primary care practices in England, de Lusignan et al. found increasing age was associated with increased odds of a positive SARS-CoV-2 test.^7^

In a meta-analysis of 32 studies, Viner et al. found that adolescents 10-14 years of age have lower susceptibility to SARS-CoV-2 infection than adults, with adolescent older than this appearing to have similar susceptibility to adults.^8^ Using a mathematical modeling approach with data from six countries (China, Italy, Japan, Singapore, Canada and South Korea), Davies et al. fitted the data available from the six countries, and estimated that the susceptibility of adolescents 10-19-year-olds is approximately half that of older adults, ages 60+ (mean susceptibility for the former is .38 and for the latter is .81), and youth in their early 20s have a susceptibility almost equal to that of older adults.^9^ However, no U.S. data was included in their model.

Regarding U.S. data on COVID-19 in adolescents, as of April 2, 2020, among 149,082 cases in all age groups for which patient age was known, only 2,572 (1.7%) of these occurred in children aged <18 years, with nearly 60% of these cases occurring in adolescents 10-17 years old. Hence at that point, adolescents accounted for just 1% of the total cases.^10^ But by Sept 15, 2020, the number of cases in adolescents 10-19 years of age had climbed to 387,000 (https://data.cdc.gov/Case-Surveillance/COVID-19-Case-Surveillance-Public-Use-Data-Profile/xigx-wn5e).^11^ The evidence in the US that adolescents are susceptible was apparent even earlier-- in June 2020--when there was an outbreak in 260 individuals at a Georgia overnight camp. In the camp’s 11-17-year-old adolescents, the attack rate was 44%, and in the 18-21-year-olds, it was 33%, where ‘attack rate’ is the number of persons with a positive test result divided by the total number of attendees in that age group (including those who did not provide testing results).^12^ Following COVID-19’s efficient spread in this youth-centric overnight setting, the CDC on July 31^st^ officially stated, “children of all ages are susceptible to SARS-CoV-2 infection and, contrary to early reports might play an important role in transmission.”^12^

Summer 2020 presented a window of opportunity to examine data on prevalence in the U.S. since adolescents and youth were on vacation and likely to mingle with others, as were adults. After months of lockdowns, there was a general easing of restrictions, with people attending parties and large gatherings, and social distancing measures were not being adhered to, leading to surges. We examined data from U.S. states experiencing surges in the number of cases to determine prevalence of COVID-19, and two other measures related to prevalence, in adolescents and youth, as compared to older adults. The comparison group are older adults since Zhang et al. found them to be most susceptible, and Davies et al. found them twice as susceptible as adolescents. Prevalence data for adolescents and youth would have important implications for school re-openings in middle school and beyond.

## METHODS

There were two criteria for inclusion of states in our study sample:

1. The state experienced a surge, defined as follows: After at least a 1-month plateau in the 7-day average of daily number of new cases, there is a a dramatic increase of at least 100% in the 7-day average number of daily new cases from the plateau 2-3 months prior, which lasts at least one month, as reported for the states in the New York Times “COVID Map and Case Count.”^13^ For example, for Missouri (https://www.nytimes.com/interactive/2020/us/missouri-coronavirus-cases.html), the case data are from August 7^th^ when there was a surge. On that day, the 7-day average number of daily new cases was 1035. On June 7^th^, 60 days prior, the 7-day daily average was 251. This is a four-fold increase (i.e., 300%) over what it was on June 7^th^. Moreover, for the months of April, May, and through June 6, the 7-day daily average of new cases had plateaued at approximately 200-300 cases per day. Also, the surge was at least one month in duration. Another example is Florida (https://www.nytimes.com/interactive/2020/us/florida-coronavirus-cases.html), where the case data are from July 19^th^. On that day the7-day daily average was 11,462. Sixty days prior, i.e., on May 19^th^, the 7-day daily average of new cases was 717. Thus, there was a 16-fold increase or 1500% increase in the 7-day daily average from May 19^th^ to July 19^th^. Also prior to that, for the months of April, May and the beginning of June, the 7-day daily average of new cases had reached a plateau, fluctuating from 578-1100 per day. Even taking the maximum or 1100, the 11,462 cases on July 19 is ten times that or a 900% increase. Also, the surge lasted at least a month from July 19, with August 19^th^ reporting a 7-day average of daily new cases at 4735.
2. The pediatric data were tabulated within distinct age brackets, and not amalgamated. Texas had over 650,000 cases, but age information was available on less than one-tenth of these, and therefore could not be included. California lumped all child data together, and also could not be included as we are excluding children under age 10 since they are not yet adolescents. We therefore considered the following six states: Florida, Tennessee, Missouri, Utah, Kansas, and South Dakota.

We accessed online tables containing COVID-19 case data from state Health Department websites when there was a surge, and tables for state population data by age group. Case data was downloaded for the summer months of July and August, between July 4^th^ and September 4^th^ (Labor Day weekend), at the time each of the states was experiencing a spike in cases. The websites are detailed in the eMethods in the Supplement.

Depending on how the data was tabulated, the case data for the six states are either for adolescents, for youth, or for both adolescents and youth combined. In South Dakota, data was tabulated by decade, and we used the 10-19-year age bracket. Tennessee had a similar age bracket of 11-20 years of age. Hence these two states provided data on adolescents. For Florida, the age brackets were 5-14, and ages 15-24 (youth, as defined by the WHO), and not for those 10-19. Therefore, we used only the latter group, the 15-24-year-olds. Similarly, For Utah, 1-14-year-olds were amalgamated, so we focused on the next age group, 15-24. Thus, Florida and Utah provided the data on youth. For Kansas, since cases were reported for 10-17 years and for 18-24 years, which consists of age bracket demarcations unlike that in any of the other states, we combined these to 10-24 years. For Missouri, we also consider 10-24-year-olds. Therefore, Kansas and Missouri provided data on adolescents and youth combined.

We calculated three measures: 1) Prevalence, 2) Percentage of Cases observed in a given age group ÷ Percentage of cases expected based on population demographics, and 3) Percentage Deviation, or [(% observed - % expected)/ % expected] x 100.

### Statistics

We performed chi-square calculations to determine whether differences between the adolescent/youth groups and the older adults were significant for the three outcome measures. Significance level was based on 2-tailed α =.05.

## RESULTS

In all states, 1) prevalence of COVID-19 for adolescents and youth was significantly greater than for older adults, p<.00001, as was 2) the ratio of observed to expected cases, p<.005 (**Table 1)**. 3) In states where the number of observed cases exceeded the expected (Utah, Missouri and Kansas), the deviation was significantly greater in adolescents/youth than in older adults, p < 0.00001. In states where observed cases were fewer than expected (Florida, Tennessee, South Dakota), the deviation for older children/youth was significantly less than that for the older adults, p < 0.00001. We now consider each developmental period separately.

**Table 1.** Prevalence-related Outcome Measures by Developmental Period and Age Bracket in US States Experiencing Spikes in COVID-19 Cases.

| | Prevalence | $\chi^2$<br>p-value | Observed ÷ Expected | $\chi^2$<br>p-value | Deviation | $\chi^2$<br>p-value |
| --- | --- | --- | --- | --- | --- | --- |
| Adolescence: South Dakota and Tennessee |  |  |  |  |  |  |
| South Dakota |  |  |  |  |  |  |
| 10-19 | 1,508/117,276<br>=<br>1.3% | $\chi^2=61.0$<br>$p<0.00001$ | 1508/1,537<br>=**98%** | $\chi^2=7.9187$<br>$p = .005$ | -29/1,537<br>=**-2%** | $\chi^2=139.0$<br>$p<0.00001$ |
| 60+ | 2234/<br>225,553=<br>0.99% |  | 2234/2,594=<br>**86 %** |  | -360/2,594=<br>**- 13.9%** |  |
| Tennessee |  |  |  |  |  |  |
| 11-20 | 14,760/<br>855,574<br>=**1.7%** | $\chi^2=1175.1$<br>$p<0.00001$ | 14,760/15,799<br>=**96%** | $\chi^2=1175.1$<br>$p=.0039$ | -1,039/15,799<br>=**-6.6%** | $\chi^2=151.0$<br>$p<0.00001$ |
| 61+ | 18,645/<br>1,577,807<br>=**1.2%** |  | 18,645/20,855<br>=**85%** |  | -2,210/<br>20,855<br>=**-10.6%** |  |
| Youth: Utah and Florida |  |  |  |  |  |  |
| Utah |  |  |  |  |  |  |
| 15-24 | 10,674/478,075=<br>**2.2%** | $\chi^2= 1548.7$<br>$p<0.00001$ | 10,674/7,034=<br>**150%** | $\chi^2= 820.9$<br>$p<0.00001$ | 3,640/ 7,034<br>=**52%** | $\chi^2=277.6$<br>$p<0.00001$ |
| 65+ | 3,686/346,281<br>=**1.1%** |  | 3,686/5,158=<br>**73%** |  | -1472/5,158=<br>**-29%** |  |
| Florida |  |  |  |  |  |  |
| 15-24 | 54,815/<br>2,555,315<br>=**2.15%** | $\chi^2=12431.5$<br>$p<0.00001$ | 54,815/41,473<br>=**132%** | $\chi^2= 6262.1$<br>$p<0.00001$ | 13,342/ 41,473<br>=**32.2%** | $\chi^2 = 14.8$<br>$p<0.0001$ |
| 65+ | 48,091/<br>4,465,169<br>=**1.08%** |  | 48,091/72,579<br>=**62%** |  | -24,488/72,579<br>=**-34%** |  |
| Adolescence and Youth: Kansas and Missouri |  |  |  |  |  |  |
| Kansas |  |  |  |  |  |  |
| 10-24 | 8,873/607,159<br>=<br>1.46% | $\chi^2= 285.5$<br>$p<0.00001$ | 8,873/7,850=<br>**113%** | $\chi^2= 135.7$<br>$p<0.00001$ | 1,023/7,850<br>=**13%** | $\chi^2=$<br>17331.3<br>$p<0.00001$ |
| 65+ | 3,979/376,116<br>=**1.06%** |  | 3,979/4,791=<br>**83%** |  | -812/4,791=<br>**-17%** |  |
| Missouri |  |  |  |  |  |  |
| 10-24 | 14,432/<br>1,192,555<br>=**1.2%** | $\chi^2= 61.6$<br>$p<0.00001$ | 14,432/<br>13,536=**105%** | $\chi^2=30.7$<br>$p<0.00001$ | 896/ 13,536<br>=**7%** | $\chi^2= 3207.1$<br>$p<0.00001$ |
| 65+ | 11,332/<br>1,033,964 =<br>**1.1%** |  | 11,332/11,731<br>=**94.6%** |  | -402/ 11,731<br>=**-3%** |  |

Legend: Chi-square statistic was for the comparison of age brackets within each state.

### Adolescence (10-19-year-olds): Data from South Dakota and Tennessee

The prevalence in adolescents was significantly greater than that in older adults (p<0.00001). This was also true for the proportion of the ratio of cases expected based on population demographics to the observed number of cases, both for South Dakota (p=0.005) and for Tennessee (p=0.0039). The third prevalence-related measure, the deviation, was significantly more negative for older adults than for adolescents (p<0.00001). This means that the ratio of [(observed cases-expected cases)/expected cases] was significantly greater for older adults than for adolescents due mainly to the fewer number of observed cases relative to expected cases in the older adults.

### Youth (15-24-year-olds): Data from Florida and Utah

In both Utah and Florida, the prevalence of COVID-19 in youth was twice what it was in older adults 65+ (p<0.00001). In both states, the ratio of observed to expected cases for youth was twice what it was for older adults (150% vs. 73% in Utah; 132% vs.62% in Florida), p<0.0001. In Utah, there were 52% more cases than expected (based on population demographics) for youth, but 29% fewer cases than expected for older adults 65+, p< 0.00001. Similarly, in Florida, whereas there were 32.2% more case than expected for youth, there were 34% fewer cases than expected for older adults 65+, p< 0.0001.

### Adolescence and Youth Combined (ages 10-24 years-old): Data from Kansas and Missouri

In both Kansas and in Missouri the data for adolescents and youth are combined. In both states, the prevalence of COVID-19 is significantly greater in the combined adolescent plus youth group than in older adults 65+ (p< 0.00001). In both states, the ratio of observed to expected cases for the combined adolescent plus youth group was significantly greater than what it was for older adults 65+ (p< 0.00001). In Kansas, there were 17% more cases than expected (based on population demographics) for youth, but 13% fewer cases than expected for older adults 65+, p< 0.00001. Similarly, in Missouri, there were 7% more cases than expected for youth, and there were 3% fewer cases than expected for older adults 65+, p< 0.0001.

## DISCUSSION

We found that prevalence of COVID-19 in adolescence was significantly greater than in older adults, and similarly for the two other prevalence-related measures. There was also a higher prevalence in youth as compared to older adults, and in adolescents and youth combined as compared to older adults. Again, the same findings held for the two other prevalence-related measures. A possible factor is that adolescents have more contacts than adults,^14^ and another factor, is that older adults, feeling vulnerable, may be more likely to adhere to masking and social distancing, which adolescents/youth may disregard. Both these factors likely act in concert to yield the pattern of results obtained.

Our findings in the six U.S. states are contrary to those of Zhang et al. in China who found that the infection rate in older adults, ages 65+, exceeded that in adolescents and youth, and to those of Wu et al., who found that of 44,672 confirmed cases of COVID in mainland China, only 1% were in adolescents ages 10-19 years of age.^15^ It is also contrary to the model of Davies et al., which estimates the susceptibility of 10-19-year-olds to be half that of older adults. One reason for the differences could be that these earlier studies were conducted when schools were closed, reducing the number of contacts adolescents and youth were exposed to, and thus the number of cases. Also, testing was less available early on, and adolescents tend to have milder cases of COVID-19 that might be missed without the availability of widespread testing. As we noted earlier, on April 2, 2020, even in the U.S., adolescents accounted for just 1% of the cases, again likely also due to the school closures and the lack of widespread testing.

A limitation of this study is that case data on state website are presumably based on people tested because they were either symptomatic, or they were exposed to someone who was thought to be infected, or they were seeking medical treatment for some other condition and the medical facility required COVID-19 testing. That still leaves some infected individuals who were asymptomatic but did not get tested because they did not fall into the latter two categories. Would their inclusion alter our results? There are two possibilities regarding asymptomatic individuals: (i) Either the number of asymptomatic infections is a constant function of the number of symptomatic ones regardless of age, as the CDC statement on June 25^th^ implied “Our best estimate right now is that for every case that’s reported, there actually are 10 other infections.”^16^ In such a scenario, the conclusions regarding our prevalence data would be unaffected since the relative proportions would remain the same. (ii) Or that the manifestation of clinical symptoms is age-dependent as Davies et al. maintain in a part of their model that deals with the clinical fraction of cases that are symptomatic vs. asymptomatic. They estimate that clinical symptoms manifest in 21% of adolescents but in 63-69% of older adults ages 60+. This would imply that there are many more asymptomatic adolescents than asymptomatic older adults: Accordingly, if asymptomatic individuals were added to our data set, our conclusions that prevalence in adolescents is significantly greater than in older adults, would be even more pronounced.

Regarding transmissibility, a large South Korean epidemiological study found that adolescents ages 10-19 were more likely to spread the virus than adults.^17^ Moreover, we know from the incident with the spread in the overnight Georgia camp that adolescents and youth can efficiently transmit the disease. Since the South Korean study and the rapid spread at the Georgia camp show that adolescents are quite capable of transmitting COVID-19 and our study shows COVID-19 to be quite prevalent in these age groups, all three studies taken together show high prevalence combined with high transmissibility.

## CONCLUSIONS

The findings of high prevalence of COVID-19 and other prevalence-related measures in adolescents and youth have implications for school re-openings. The age groups 10-19 and 15-24 are typically students in middle school, high school, college, and the beginning of graduate/professional school. Our findings would make an argument against school re-openings in the near future. In places, where schools have nevertheless reopened, the high prevalence of COVID-19 highlights the necessity of students, faculty and staff wearing masks, social distancing, and washing hands regularly.

## Data Availability

All data is available at the listed URLs.

https://floridahealthcovid19.gov/

https://www.tn.gov/content/tn/health/cedep/ncov/data.html

http://mophep.maps.arcgis.com/apps/MapSeries/index.html?appid=8e01a5d8d8bd4b4f85add006f9e14a9d

https://www.coronavirus.kdheks.gov/160/COVID-19-in-Kansas

https://coronavirus.utah.gov/case-counts/

https://doh.sd.gov/news/coronavirus.aspx

http://edr.state.fl.us/Content/populationdemographics/data/Pop_Census_Day.pdf

https://www.tn.gov/content/dam/tn/health/documents/population/TN-Population-by-AgeGrp-Sex-Race-Ethnicity-2019.pdf

https://healthapps.dhss.mo.gov/MoPhims/QueryBuilder?qbc=PNM&q=1&m=1

https://gardner.utah.edu/wp-content/uploads/State-of-Utah-Demographic-Profile-2010-2018.pdf

## Notes

### Competing Interest Statement

The authors have declared no competing interest.

### Funding Statement

No funding was secured for this study.

### Author Declarations

Study is exempt. Data are de-identified data from the six states' Department of Health websites.

## REFERENCES

1. Dong, Y., Mo, X., Hu, Y., Qi, X., Jiang, F., Jiang, Z. and Tong, S. Pediatrics June 2020, 145 (6) e20200702. DOI: https://doi.org/10.1542/peds.2020-0702

2. Lu X, Zhang L, Du H, Zhang J, Li YY, Qu J, Zhang W, Wang Y, Bao S, Li Y, Wu C, Liu H, Liu D, Shao J, Peng X, Yang Y, Liu Z, Xiang Y, Zhang F, Silva RM, Pinkerton KE, Shen K, Xiao H, Xu S, Wong GWK; Chinese Pediatric Novel Coronavirus Study Team. SARS-CoV-2 Infection in Children. N Engl J Med. 2020 Apr 23;382(17):1663–1665. doi: 10.1056/NEJMc2005073.

3. Bi, Q., Wu, Y., Mei, S., Ye, C., Zou, X., et al. Epidemiology and transmission of COVID-19 in 391 cases and 1286 of their close contacts in Shenzhen, China: a retrospective cohort study. Lancet Infect. Dis. (2020); https://doi.org/10.1016/S1473-3099(20)30287-5

4. Zhang, J., Litvinova, M., Liang, Y., Yan Wang, Y., Wei Wang, W. et al. Changes in contact patterns shape the dynamics of the COVID-19 outbreak in China. Science, 368, 1481–1486 (2020). DOI: 10.1126/science.abb8001

5. Geliebter, A., Rumain, B., & Schneiderman, M. Re: Changes in contact patterns shape the dynamics of the COVID-19 outbreak in China. Science (2020). Published online 26 June 2020. https://science.sciencemag.org/content/early/2020/05/04/science.abb8001/tab-e-letters

6. Kuchar E, Załęski A, Wronowski M, et al. Children were less frequently infected with SARS-CoV-2 than adults during 2020 COVID-19 pandemic in Warsaw, Poland [published online ahead of print, 2020 Sep 28]. Eur J Clin Microbiol Infect Dis. 2020;1–7. doi:10.1007/s10096-020-04038-9

7. de Lusignan S, Dorward J, Correa A, Jones N, Akinyemi O, Amirthalingam G, Andrews N, Byford R, Dabrera G, Elliot A, Ellis J, Ferreira F, Lopez Bernal J, Okusi C, Ramsay M, Sherlock J, Smith G, Williams J, Howsam G, Zambon M, Joy M, Hobbs FDR. Risk factors for SARS-CoV-2 among patients in the Oxford Royal College of General Practitioners Research and Surveillance Centre primary care network: a cross-sectional study. Lancet Infect Dis. 2020 Sep;20(9):1034–1042. doi: 10.1016/S1473-3099(20)30371-6.

8. Viner, RM, Mytton, OT, Bonell, C., Melendez-Torres, GJ, Ward, JL, Hudson, L., Waddington, C., Thomas, J., Russell, S., van der Klis, F., Koiral, A., Ladhani, S., Panovska-Griffiths, J., Davies, NG, Booy, R., Eggo, R. Susceptibility to and transmission of COVID-19 amongst children and adolescents compared with adults: a systematic review and meta-analysis. medRxiv 2020.05.20.20108126; doi: https://doi.org/10.1101/2020.05.20.20108126

9. Davies, N.G., Klepac, P., Liu, Y., Prem, K., Jit, M., et al. Age-dependent effects in the transmission and control of COVID-19 epidemics. Nat Med (2020). Published 16 June 2020. https://doi.org/10.1038/s41591-020-0962-9

10. CDC. COVID-19 Response Team. Coronavirus Disease 2019 in Children - United States, February 12-April 2, 2020. MMWR Morb Mortal Wkly Rep. 2020 Apr 10;69(14):422–426. doi: 10.15585/mmwr.mm6914e4. PMID: 32271728; PMCID: PMC7147903

11. CDC. COVID-19 Case Surveillance Public Use Data Profile. https://data.cdc.gov/Case-Surveillance/COVID-19-Case-Surveillance-Public-Use-Data-Profile/xigx-wn5e [Retrieved October 09, 2020].

12. CDC. SARS-CoV-2 Transmission and Infection Among Attendees of an Overnight Camp — Georgia, June 2020. Morbidity and Mortality Weekly Report (MMWR). Early Release July 31, 2020. (https://www.cdc.gov/mmwr/volumes/69/wr/mm6931e1.htm?s_cid=mm6931e1_x

13. https://www.nytimes.com/interactive/2020/us/coronavirus-us-cases.html

14. Mossong, J. et al. Social contacts and mixing patterns relevant to the spread of infectious diseases. PLoS Med. 5, e74 (2008).

15. Wu, Z & McGoogan, JM. Characteristics of and important lessons from the coronavirus disease 2019 (COVID-19) outbreak in China: summary of a report of 72 314 cases from the Chinese center for disease control and prevention. JAMA 2020. [Retrieved October 8, 2020].

16. Redfield, Robert. (June 25, 2020). CDC says COVID-19 cases in U.S. may be 10 times higher than reported. NBC News. https://www.aol.com/article/news/2020/06/25/cdc-says-covid-19-cases-in-us-may-be-10-times-higher-than-reported/24537120/

17. Park YJ, Choe YJ, Park O, Park SY, Kim YM, Kim J, et al. Contact tracing during coronavirus disease outbreak, South Korea, 2020. Emerg Infect Dis. 2020 Oct [Retrieved July 19, 2020]. https://doi.org/10.3201/eid2610.201315

