## Supplemental Information for "Prevalence of COVID-19 in Adolescents and Youth Compared with Older Adults in States Experiencing Surges"

**SUPPLEMENTARY INFORMATION**

**eMethods**

This supplementary material is provided by the authors to give readers additional source information.

Case data state websites:

Florida: <https://floridahealthcovid19.gov/> [retrieved July 19, 2020];

Tennessee: <https://www.tn.gov/content/tn/health/cedep/ncov/data.html> [Retrieved August 12, 2020],

Missouri<http://mophep.maps.arcgis.com/apps/MapSeries/index.html?appid=8e01a5d8d8bd4b4f85add006f9e14a9d> [Retrieved August 7, 2020]:

Kansas: https:// www.coronavirus.kdheks.gov/160/COVID-19-in-Kansas [Retrieved Aug 23, 2020];

Utah: [https://coronavirus.utah.gov/case-counts/[Retrieved](https://coronavirus.utah.gov/case-counts/%5bRetrieved) August 18, 2020];

South Dakota <https://doh.sd.gov/news/coronavirus.aspx> [Retrieved Sept. 4, 2020].

Demographic data by age and by state websites:

Florida: <http://edr.state.fl.us/Content/populationdemographics/data/Pop_Census_Day.pdf>,

Tennessee: <https://www.tn.gov/content/dam/tn/health/documents/population/TN-Population-by-AgeGrp-Sex-Race-Ethnicity-2019.pdf>**;**

Missouri: <https://healthapps.dhss.mo.gov/MoPhims/QueryBuilder?qbc=PNM&q=1&m=1>;

Utah: <https://gardner.utah.edu/wp-content/uploads/State-of-Utah-Demographic-Profile-2010-2018.pdf> ; Kansas: <https://ipsr.ku.edu/sdc/region.php?area=Kansas&tab=1#TOTPOP>;

South Dakota: <https://www.sdstate.edu/sociology-rural-studies/census-data-center/population-change>

New York Times Websites indicating surges by state:

<https://www.nytimes.com/interactive/2020/us/florida-coronavirus-cases.html>

<https://www.nytimes.com/interactive/2020/us/tennessee-coronavirus-cases.html>

<https://www.nytimes.com/interactive/2020/us/missouri-coronavirus-cases.html>

<https://www.nytimes.com/interactive/2020/us/kansas-coronavirus-cases.html>

<https://www.nytimes.com/interactive/2020/us/utah-coronavirus-cases.html>

<https://www.nytimes.com/interactive/2020/us/south-dakota-coronavirus-cases.html>
